# Good Intentions but bad outcomes – Communal-bowl hand-rinsing before meals transmits pathogens and diseases: a systematic review

**DOI:** 10.1101/2024.08.19.24311865

**Authors:** Evans Atiah Asamane, Katie Scandrett, Justin A. Aunger, Alexander Browne, Anoushka Ramkumar, Cheick Sidibe, Youssouf Diarra, Mariam Yazdani, Ousmane Toure, Richard Lilford, Semira Manaseki-Holland

## Abstract

**Background:** Communal Bowl-Hand Rinsing (CB-HR), refers to more than one person washing their hands in one shared container of water, without soap and without changing the water between individuals, before meals. CB-HR has persisted in Africa despite decades of handwashing education during communicable disease epidemics, including Covid-19. We systematically reviewed the literature to provide a better understanding of the spread and motivations for the practice, its association with disease or pathogen transmission, and targeted interventions.

**Methods:** Several electronic databases and grey literature were searched. We extracted data and appraised their methodological rigour using the Mixed Methods Appraisal Tool (MMAT). A narrative synthesis and Forest Plot were used to summarise the data.

**Findings:** Fifteen studies (all from 10 Sub-Saharan Africa (SSA) countries) were identified from 10,711 records. Study settings were schools (n=4), funerals (n=1), and households (n=10). Four case-control studies reported increased odds of cholera (Adjusted-OR=6.50;95%CI,2.30, 18.11), dysentery (at households Adjusted-OR=10.52(95%CI,2.81, 39.0), and at public gatherings Adjusted-OR=2.92(95%CI,1.24,7.21)), diarrhoea (Adjusted-OR=2.89;95%CI,1.33, 6.39), Hepatitis E virus risk (Adjusted-OR=1.90; 95%CI,1.07, 3.38), and one found a lower height-for-age z-score in babies whose families perform CB-HR. A cross-sectional study reported odds of cysticercosis (Adjusted-OR=3.8;95%CI,2.5, 5.9). Two cross-sectional studies conducted laboratory water and/or hand-swab microbiology investigations, demonstrating pathogen transmission from CB-HR. No intervention studies were found.

**Conclusion:** Research on CB-HR was sparse. CH-BR is widely practised in SSA and spreads diseases. However, there is a need for the development and evaluation of culturally sensitive interventions to address this practice in Africa.

The review was not funded but the authors were funded by various grants.

**Key Messages:** *What is already known on this topic:* Gastrointestinal diseases are an important cause of disease and death, especially in young children. The risk of Gastrointestinal diseases can be reduced by hand hygiene. Extremely limited primary research has been conducted into the health effects of Communal Hand Rinsing (CB-HR) in a shared bowl of water before meals. CB-HR simply refers to the practice before eating where more than one person wash/rinse their hands in one shared container of water, without soap and without changing the water between individuals. No systematic reviews have been conducted on this topic.

*What this study adds:* This systematic review collates all available research on the prevalence, motivators, and disease transmission of pathogens following CB-HR, thus providing important new insights about this practice. CB-HR is a common practice and found to be a risk factor for the spread of communicable diseases such as diarrhoea, cholera, and dysentery.

*How this study might affect research, practice or policy:* Further research is urgently needed to develop culturally sensitive interventions to reduce the risk of disease associated with CB-HR.

## Introduction

Effective handwashing has been shown to be the most successful and cost-effective method to reduce the transmission of numerous communicable diseases.^1^ Systematic reviews have demonstrated that Water, Sanitation and Hygiene (WASH) interventions including handwashing with soap and water can reduce the risk of diarrheal diseases by 30%,^2^ respiratory diseases by 16%,^3^ and child mortality by 17%.^4^ Most recently, frequent and effective handwashing was encouraged as key, alongside other measures, to combat the ongoing global Coronavirus Disease 2019 (COVID-19) pandemic.^1,3^

Handwashing using soap and running water, is not practiced equally worldwide.^5^ Cultural beliefs shape methods of handwashing. A common but under-researched handwashing practice carried out in many countries in sub-Saharan Africa is Communal Bowl-Hand Rinsing before meals (CB-HR). CB-HR refers to the practice of a group of people (two or more people) washing their hands in one container of water without soap and without changing the water between individuals. The practice seems to have persisted in many African countries for years including countries that have experienced devastating epidemics of cholera, Ebola and COVID-19.^1,6–8^ CB-HR practice poses a risk, as the contaminated water acts as a route for diseases and pathogen transmission.^6,9,10^

Given the potential impact of CB-HR before meals, we aim to provide a comprehensive review of the current literature concerning CB-HR by analysing and reporting on prevalence, risk and motivations of CB-HR practice. In addition, we will also explore current interventions to specifically target CB-HR, globally.

## Methods

### Search strategy and selection criteria

We followed the Preferred Reporting Items for Systematic Reviews and Meta-Analyses (PRISMA) guidelines, with protocol registered with PROSPRO ID: CRD42021277767.^11^

#### Literature Search

A systematic literature search of the following databases was carried out: Applied Social Sciences Index & Abstracts (ASSIA), Cumulative Index of Nursing and Allied Health Literature (CINAHL), the Cochrane Library, EMBASE, EThOS, MEDLINE, PsychInfo and Scopus. Grey literature searches were conducted through the search engines Google and Web of Science. Details of these search terms can be found In Appendix A. Additionally, reference lists of eligible studies were searched for other relevant papers. Once studies were identified via database searching and reference scanning. Citation tracking was performed for all included studies using Google Scholar to identify any eligible studies that may have cited the included ones. See Appendix A for details of the search strategy used for the various databases. The identified studies were stored and managed using endnote reference management software. Duplicates were automatically removed using EndNote before the selection phase. Searches were updated and expanded to include studies outside Africa. Finally, selected experts were consulted for their knowledge of publicised studies.

#### Eligibility Criteria

Studies were eligible for inclusion if they were observational, experimental, or qualitative in design and were conducted in any population where CB-HR was practised. All included studies were published in English or French. Studies that analysed food, household, or other hygiene practices as outcomes without exploring any aspect of CB-HR were excluded. A table detailing the inclusion and exclusion criteria can be found in Appendix B.

#### Study Selection, Quality Assessment and Data Extraction

Two reviewers (AB and AR) selected studies independently through a two-stage process: firstly, through title and abstract screening, then full-text screening. We contacted two authors to clarify aspects of the study methodology, outcomes, population or study setting.

The Mixed Methods Appraisal Tool (MMAT) Version 2018 was used to assess the methodological quality of included studies.^12^ MMAT tool has been frequently tested for reliability in the literature and it is a commonly used tool for systematic reviews with mixed methods studies.^13^ Two reviewers (EAA and JAA) assessed the studies independently, following the tool guidelines.

Afterwards, data was extracted from all the included studies using a bespoke data collection form, which was piloted on three included studies and modified before use to ensure all relevant data was collected. Using this form, we collected study descriptive and quality data (Table 1). The pre-defined CB-HR outcomes extracted are listed in Table 1.

**Table 1.**

| Author (Year)<br>City, Country<br>Year | Study design | Sample Size and<br>Recruitment/Sampling | Study summary and<br>Socio-demographic<br>information | Analysis<br>CB-HR relevant results | Quality of study<br>MMAT quality score<br>(0 lowest and 5<br>highest)* (MMAT<br>2018) |
| --- | --- | --- | --- | --- | --- |
| Howard et al. <sup>6</sup><br>2010.<br>Kitgum, Uganda | Matched Case-control | 257 total participants<br>112 cases (patients with jaundice (scleral icterus) and serological evidence of HEV infection) recruited by consecutive referral from the adjacent health centre.<br>145 controls (persons with no symptoms/diagnosis of those diseases)- recruited from the residents of villages in this area and a camp for internally displaced persons<br>Sampling technique: not reported | Investigating risks for an outbreak of HEV in urban and rural areas including in an IDP camp in the area.<br>**SDC data/Matched for:**<br>-Age group - < 5, 5–14, 15–45, and > 45 years; and<br>-Location of primary residence (in the past 8 weeks)<br>No SDC data provided. | **Analysis:** Inferential analysis<br>**Odds ratio for disease transmission:**<br>OR (95% CI): 2.17 (1.25,3.79);P = 0.006<br>AOR (95% CI): 1.90 (1.07,3.38); P= 0.028<br>CB-HR was a 'risk factor' for HEV.<br>Authors comment (no data) that CB-HR is common in the setting, especially in gatherings such as funerals | 3 |
| Maponga et al. <sup>7</sup><br>2013<br>Kadoma,<br>Zimbabwe | Case-control | 218 total participants<br>109 cases (<5 years with diarrhoea) selected via multi-stage sampling from clinics and patient line lists. | Investigating the risk factors for contracting watery diarrhoea in children < 5 years in two townships (urban).<br>**SDC data:** | **Analysis:** Descriptive and Inferential statistical analysis<br>**Odds ratio for disease transmission:**<br>OR (95% CI): 2.95 (91.47,5.89)<br>AOR (95% CI): 2.89 (1.33, 6.28)<br>CB-HR was a 'risk factor' for diarrhoea seen | 4 |
|  |  | 109 controls (<5 years without diarrhoea)-selected from a household 10 houses away from a case.<br>Sampling technique: Multistage | Cases:<br>Male: 55.0%<br>Caregiver education primary or below: 11.0%<br>Occupation of caregiver: Housewife (86%)<br>Controls:<br>Male: 51.0%<br>Caregiver education: primary or below: 15.0%<br>Occupation of caregiver: Housewife (87%) | at clinics. |  |
| Oyugi et al. <sup>8</sup><br>2017<br>Rongo and Ndhiwa, Kenya | Matched Case-Control | 156 total participants<br>52 cases (with watery diarrhoea) Systematic random sampling using patient list was used, and selected cases traced to their household in the community.<br>104 controls (No watery diarrhoea) randomly spinning a bottle while still at the home of a case, the nearest home to the case in the direction of the bottle was selected.<br>Sampling technique: Systematic random sampling | Investigation of factors associated with Cholera outbreak in two urban counties.<br>**SDC data/Matched for:**<br>Age group (2-5, > 5-15, > 15-25, > 25-39, > 39 years)<br>Village of residence<br>SDC Cases:<br>Male: 38%<br>Christian: 50%<br>No education: 8%<br>SDC Control:<br>Male: 39%<br>Christian: 55%<br>No education: 7% | **Analysis:** Descriptive and Inferential statistical analysis<br>**Odds ratio for disease transmission for those reporting CB-HR:**<br>AOR (95% CI):6.5(2.30, 18.11).<br>CB-HR was a 'risk factor' for cholera seen at clinics. | 4 |
| Midzi et al.<sup>21</sup><br>2000<br>Nyaure Ward, Zimbabwe | Case-control | 104 total participants<br>52 cases (every other case with bloody diarrhoea presented to a local clinic)<br>52 controls (with no bloody diarrhoea from the case's family of neighbour <200m from case's home)<br>Sampling/recruitment: No reported | Investigating the risk factors associated with Shigella dysenteriae 1 in an urban community.<br>**SDC data:**<br>Median age cases = 17 years (Q1 = 8, Q3 = 30)<br>Median age controls = 19 years (Q1 = 7, Q3 = 28) | **Analysis:** Descriptive analysis.<br>**Odds ratio for disease transmission**<br>CB-HR was associated with contracting Shigella dysenteriae 1 (dysentery) in different settings:<br>Public gatherings:<br>OR (95% CI): 8.47 (2.43, 31.33)<br>AOR (95% CI): 2.90 (1.24, 7.21)<br>In the home:<br>OR (95% CI): 60.43 (15.73, 256.00)<br>AOR (95% CI): 10.52 (2.81, 39.00)<br>**Motivations/reasons for CB-HR**<br>Authors speculate that washing hands in a common bowl of water at public gatherings like funerals is a common/usual practice. | 4 |
| Rukambile et al.<sup>22</sup><br>2020<br>Sanza, Majiri and Iwondo wards, Tanzania | Cohort | 493 pair of participants<br>Sampling technique: Two stage sampling.<br>- Households with children <12 months old.<br>- Random selection of additional households through a lottery draw using identification numbers.<br>493 pairs of mothers/caregivers and children 12-24m in 493 households, randomly sampled out of 711 households | Investigating factors associated with diarrhoea and z-score height in a rural community with 24m follow-up of children<br>**SDC data:**<br>Male: 47.7%<br>Mean child age (SD): 32 (5.3) months<br>Mean Maternal age (SD): 32 (7.5) years<br>Maternal primary education level: 71.4% | **Analysis:** Inferential analysis,<br>**Risk to outcomes:**<br>Height-for-age z-score: Univariate coefficient for running water vs same container handwashing 0.24 SE=0.10, P=0.009<br>Multivariate coefficient 0.27 SE=0.10, P=0.007.<br>Diarrhoea: Univariate coefficient for running water vs same container handwashing -0.02, P=0.78<br>(no frequency or 95%CI given) | 4 |
| Mwanjali et al.<sup>24</sup><br>2013<br>Mbozi district, Tanzania | Cross-sectional | 13 randomly selected villages in a district of which 900 households were randomly sampled. One adult 16-60 year randomly sampled per household (70 households dropped out, 830 total participants included) | Investigating the prevalence and risk factors of human *T. solium* infections in rural villages. Investigating symptoms, lab analysis EITB assay (faecal) and Ag-ELISA assay (serum), CT brain<br>**SDC data:**<br>Mean age (SD): 37.9(11.3) years<br>Male: 57%<br>Primary education level: 77.1%<br>Christian: 82.4%<br>Farming occupation: 85.4% | **Analysis:** Inferential analysis<br>**Scale of practice**<br>451 out of 830 (54.3%) participants practiced CB-HR without soap<br>**Odds ratio for risk of disease:**<br>AOR (95% CI): 3.8 (2.5, 5.9), P = 0.0001.<br>CB-HR was associated with increased odds of Ag-active ELISA +ve human cysticercosis.<br>No counts or ratios given. | 5 |
| Moabi<sup>9</sup><br>2016<br>Mangaung South Africa | Cross-sectional | 42 funerals purposively sampled, 126 water samples of before and after CB-HR (3 samples per funeral – 1 of the tap in the yard, 1 of the water bowl before CB-HR, and 1 after all CB-HR washes taken place) | Investigating the microbial quality of CB-HR water in funerals in an urban municipality.<br>No SDC data provided | **Analysis:** Descriptive analysis<br>**Microbial content of water samples:**<br>Compliance of bowl water using safe microbial South African National Standard (lower value indicates less safe water)<br>Compliance before CB-HR:<br>Aerobic bacteria – 95.2%<br>Coliforms – 78.6%<br>Escherichia coli – 88.0%<br>Compliance after CB-HR:<br>Aerobic bacteria – 45.2%<br>Coliforms – 2.4% | 2 |
|  |  |  |  | Escherichia coli – 47.6%<br>**Description:** Mourners often used the same bowl to rinse hands before going to the cemetery and after returning, before eating.<br>**Motivations/reasons for CB-HR**<br>**Authors without data speculate that** CB-HR attributed to religious and spiritual reasons and purification (to wash bad omens away after returning from the cemetery). |  |
| Tetteh-Quarcoo et al.<sup>10</sup><br>2016<br>Accra, Ghana | Cross-sectional | 6 preschools purposively sampled. Either part of Ghana School feeding programme or private schools providing meals for children.<br>1 water sample from the main, 1 from soapy bowl water and 1 from rinse water, and 3 randomly selected pupils from each school were swabbed before and after washing in communal bowl. | Investigating the microbial content of “bowl water” used for CB-HR in preschools in urban areas<br>6 Preschools,<br>**SDC data:**<br>Private schools: 5<br>Government feeding program: 1 | **Analysis:** Descriptive analysis.<br>**Microbial content and hand swabs:**<br>Before CB-HR hand-swabs only +ve were: Staphylococcus (33%) and A.niger (5%).<br>After CB-HR hand-swabs +ve were: Staphylococcus (41.6%), Salmonella (25%), E.coli (22%), Citrobacter (16%), K.oxytoca (11%), Klebsiella (11%), A.niger (5%).<br>All bacteria were resistant to 3 common antibiotics. | 2 |
| Melariri et al.<sup>19</sup><br>2019<br>Nelson Mandela Bay, South Africa | Cross-sectional | 46 ECD centres (ECD Training Centres) were purposively sampled which had a total enrolment of 2607 children and caregivers (2435 children, 172 caregivers) | Investigating Water Sanitation and Hygiene (WASH) status and practices in ECD centres in low socio-economic areas (urban)<br>**SDC data for children in ECDs:**<br>Age range of children: 6 months – 6 years = 2,435.<br>Male: 46.6% | **Analysis:** Descriptive analysis<br>**Scale of practice (Observed)**<br>36 out of 46 (79%) ECD centres used CB-HR.<br>**Motivations/reasons for CB-HR**<br>(not clear if this is speculation from authors or actual questionnaire replies):<br>“At 36(79%) of CDs, children collectively shared plastic bowls for handwashing due to absence of running water.”<br>A pie-chart show in some ECDs >30 children shared one bowl. | 3 |
| Mwapasa et al.<sup>20</sup><br>2022<br>Blantyre, Malawi | Cross-sectional | 10 pre-school ECDs randomly selected from 900 ECDs (882= 849 children and 33 caregivers)<br>261 microbial samples (water and hand-swabs) collected | Investigating hygiene practices in ECDs in low socio-economic urban areas.<br>**SDC data:**<br>Child caregivers at ECD centres:<br>Female: 100%<br>Primary education level: 70%<br>Children:<br>Age range: 2-6 years.<br>Average ratio of caregiver to children (1:77). | **Analysis:** Mixed methods-Quantitative, descriptive statistical analysis. Qualitative, constant comparison analysis<br>**Scale of practice:**<br>94 out of 849 (11.1%) children practiced CB-HR without soap across ECDs in the sample observed.<br>“On some occasions, children were observed drinking from the CB-HR container” | 3 |
| Appiah-Brempong et al.<sup>23</sup><br>2018<br>Ejisu-Juaben, Ghana | Cross-sectional | 37 junior high schools (5661 students) selected via stratified random sampling from 80 schools in the municipality. | Investigating handwashing practices and hygiene facilities in schools in an urban municipality.<br>**SDC data:**<br>37 mixed gender public schools<br>Mean number of students per school: 153, SD: 81.9<br>68% of schools used soap | **Analysis:** Descriptive analysis<br>**Scale of practice and speculated reasons for CB-HR practice**<br>12 out of 37 (33%) schools practiced CB-HR; 5 schools (13.5%) had functional handwashing facilities.<br>**Motivations/reasons for CB-HR**<br>**Authors speculate reasons for CH-BR practice** being related to a lack of handwashing facilities or water supply, even though government funds for these were available for some schools. | 5 |
| Ehiri et al.<sup>16</sup><br>2001<br>Imo state, (Owerri, okigwe, orlu zones), Nigeria | Cross-Sectional | 120 total Households (one child per household) selected via multistage sampling technique from 601 households | Investigating microbial contamination and critical control points (HACCP) in the preparation and handling of complementary foods in both urban and rural areas.<br>Observations and questionnaire.<br>**SDC data:**<br>Mean age: 15 months<br>Male: 35%<br>Mean members per household: 7 | **Analysis:** Little reporting for CB-HR as detailed below with no other data.<br>**Scale of CB-HR practice;** in text it is stated: "In 65 households where children were fed during household mealtimes, all household members washed their hands (without soap) in one bowl of water before eating." | 4 |
| Dancesco et al. <sup>15</sup><br>2005<br>Tiaha, Côte d'Ivoire | Cross-sectional | 416 total participants<br>Sampling: Not reported | Intestinal parasitology examinations (using perianal swabs and stools) in a rural fishing community.<br>**SDC data:**<br>Male: 55%<br>7% aged 6 months-3 years,<br>82% aged 4-15 years;<br>11% adults | **Analysis:** Little reporting for CB-HR.<br>**Scale of CB-HR practice:**<br>It is speculated in text by authors without data that the prevalent practice of CB-HR in homes and schools in the area would be responsible for high rates of intestinal parasites in children.<br>**Motivations/reasons for CB-HR**<br>Authors speculate without data that reasons/motivations for CH-BR is the lack of running water, even if soap is readily available. | 2 |
| Schmitt et al. <sup>17</sup><br>1997<br>City not reported, Zambia | Cross-sectional | 17 total Households with a diarrhoea case, purposively selected using protocol: diarrhoea cases from a diarrhoea clinic serving 2 townships or also asking individuals at one community well in one of the 2 townships. | HACCP analysis of food preparation in homes with a diarrhoea case.<br>**SDC data:**<br>Two urban areas<br>Average household membership: 8 people<br>Average number of children <5 years in each household: 2<br>No other SDC data provided | **Analysis:** Little reporting for CB-HR as detailed below with no data<br>**Scale of CB-HR practice**<br>In text the authors state: "further source of hand contamination was the customary practice of rinsing hands in a common bowl of water prior to eating.... populations of thermotolerant coliforms were found in samples of leftover foods. This observation provides evidence of post-cooking contamination which could have occurred during serving or sharing portions of the implicated foods during eating." | 1 |
| Badowski et al. <sup>18</sup><br>2011<br>Dar Es Salaam, | Cross-sectional qualitative/observational | 13 total female heads of households from 2 peri-urban communities were purposively selected for similarity of their | Using modified PhotoVoice method to capture daily activities of mothers in peri-urban areas. | **Analysis:** Qualitative, PhotoVoice<br>**Scale of practice**<br>Household interviews made 30 references to | 3 |
| Tanzania |  | socioeconomic characteristics<br>(127 photographs collected] | **SDC data:**<br>Women's aged 18–50 years<br>No other SDC given. | handwashing (including CB-HR) Reference to handwashing (no mention of soap) in photos/ interviews were made by 9 households. Details are vague but references to dipping hands in communal bowls before eating. CB-HR described as habitual before handling and consuming food.<br>**Motivations/reasons for CB-HR**<br>CB-HR practice habitual as previous droughts and scarcity of water compelled households to practice CB-HR to save water which had persisted even at times when water was plentiful. |  |

**Abbreviations:** Ag-ELISA – Antigen - enzyme-linked immunosorbent assay; ECD– Early Childhood Development; SDC – Sociodemographic Data. \* MMAT Tool (version 2018) guidance notes advises against calculating an overall score from the ratings of each criterion but rather to present the details of the ratings per each criterion to allow a more informed interpretation of the quality of the included studies. We have presented the MMAT tool table for all included studies in Appendix C.

#### Forest Plot and Narrative Analysis

We presented results visually on a Forest plot for studies reporting effect measures relating CB-HR exposure to disease (Figure 3).

A narrative synthesis was conducted based on the guidance of Popay and colleagues^14^; this was appropriate given the lack of interventional studies and the heterogeneity of included studies.

## Results

In line with our eligibility criteria, 15 eligible studies were included (Figure 1). The excluded studies were mainly studies without a mention of CB-HR practice. All included 15 studies were published from 1997-2022.

**Figure 1:**
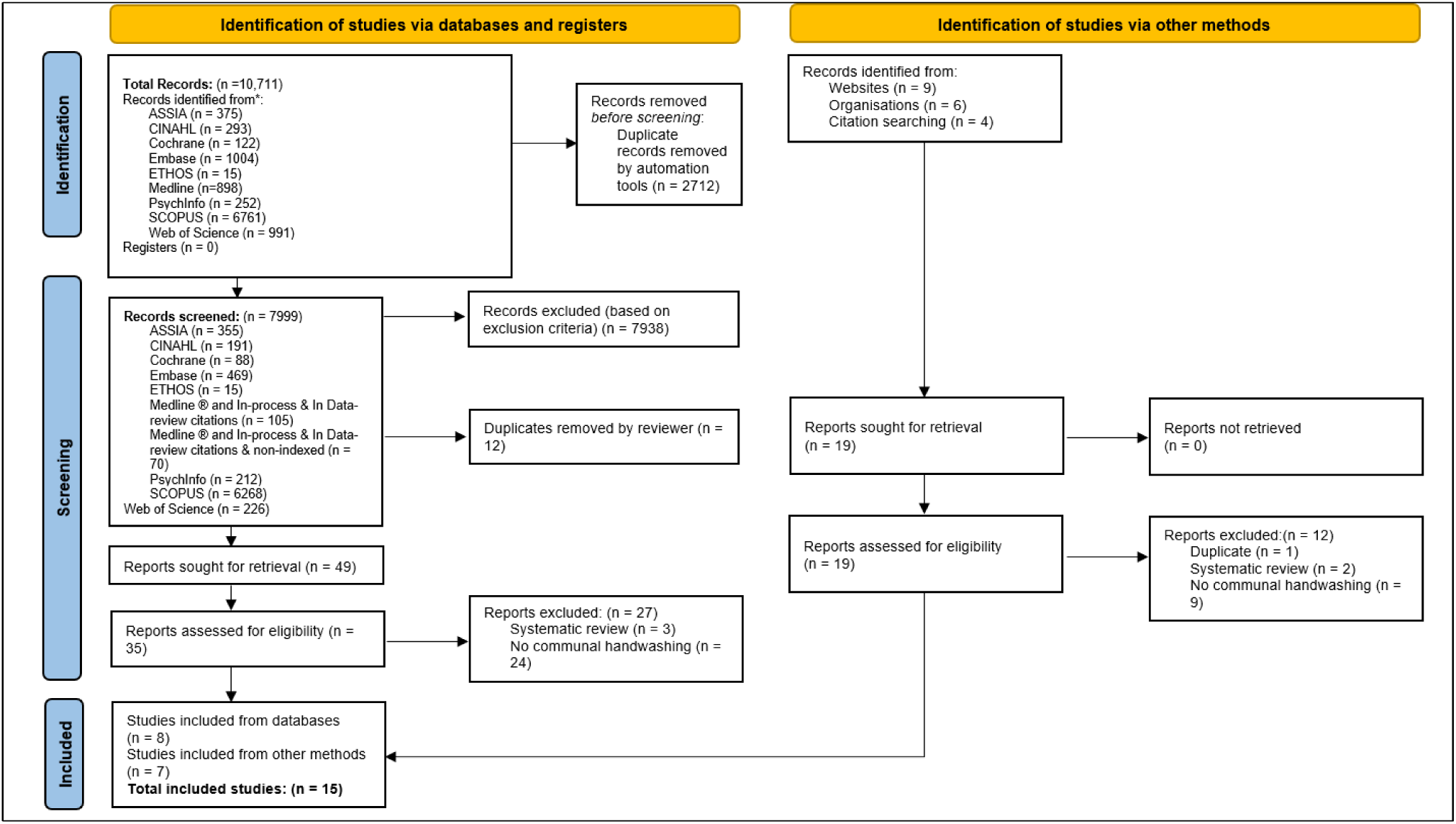
Flow Diagram illustrating the searches and included studies.

Three studies made mention of CB-HR practice in their study population in the text of the paper with little to no data presented.^15–17^ We decided to include these given the small number of studies and as an indication of CB-HR being practiced in the study population to help the assessment of the spread of the practice.

### Quality of studies

Using the MMAT 2018 tool for quality assessment with a score of 5 being the highest and 0 the lowest, most studies were rated 3 and below. One study scored 1,^17^ three studies scored 2,^9,10,15^ four studies scored 3,^6,18–20^ five studies scored 4,^7,8,16,21,22^ and the remaining two studies scored 5.^23,24^ See details in Appendix C.

### Description of CB-HR

There are two main types of CB-HR. One is a pre-prandial ritual whereby several people rinse their hands in one big bowl of water simultaneously, and rub/rinse their hands before eating. The second form of CB-HR is where each member of the household individually and sequentially dips/rinses in a common bowl before eating

### Location/Setting of included studies

All studies took place within ten Sub-Saharan African countries. Of the 15 included studies, five were conducted in eastern Africa (Kenya, Tanzania and Uganda),^6,8,18,22,24^ six in southern Africa (South Africa, Zambia, Malawi and Zimbabwe)^7,9,17,19–21^ and four in western Africa (Nigeria, Ghana and Côte d’Ivoire).^10,15,16,23^ This is visually represented in Figure 2. Studies were carried out in many settings within these countries. Five studies involved entire states or regions of the country^6,16,21–23^ and seven in major metropolitan areas that were either national or state/district capital cities.^7–10,18–20^ One study was conducted in two townships near a large city^17^ and two in smaller rural towns/villages.^15,24^ Within these settings, four studies were in schools, preschools or Early Development Centres (ECDs), ^10,19,20,23^ one at funerals^9^, five in households^15,16,18,22,24^, and five were of clinic patients and community member controls.^4,6–8,21^

**Figure 2:**
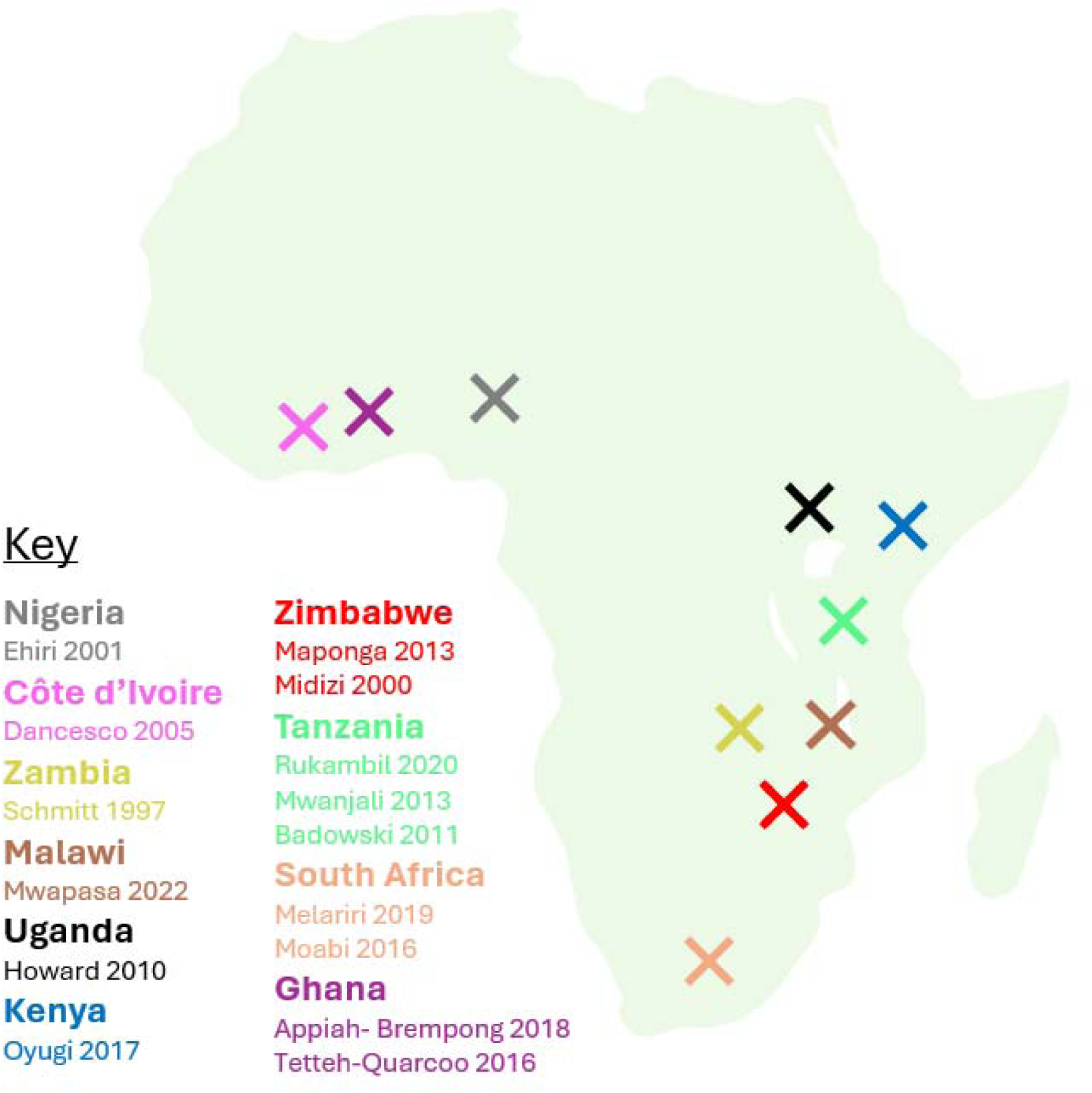
Map showing the locations of where the included studies were conducted.

Demographics of included studiesFour studies^10,19,20,23^ were conducted exclusively in school or preschool aged children, of which two also included their teachers.^19,20^ Mwapasa et al.^20^ and Melariri et al. ^19^collected data from children attending ECD centres as well as the teachers (ECD caregivers) operating them. One study was conducted with children exclusively under-5 years old.^7^ Rukambile^22^ studied pairs of mothers/caregivers and children.

The remaining studies that reported demographics had adolescents or adults as participants. Full sociodemographic information is presented in Table 1. However, participants’ socio-demographic data were often not detailed or not reported as indicated in Table 1.

### Study designs and Outcomes of included studies

Four studies were case-control design,^6–8,21^ one a cohort study,^22^ nine were cross-sectional designs,^9,10,15–17,19,20,23,24^ and one a qualitative Photovoice design.^18^

Outcomes of interest varied, with five of the studies primarily looking at risk factors for certain diseases, with CB-HR being one of these factors.^6–8,21,24^ One also investigated risk factors associated with height-for-age z-score in young children.^22^ Four studies included outcomes specifically relating to the availability, quality and use of handwashing facilities at schools or ECD centres.^10,19,20,23^ Two studies examined the microbiology of hand-swabs or of “communal-bowl water”.^9,10^

### Disease transmission risk

Four case-control studies reported increased odds of various diseases or infections for those who engaged in CB-HW compared to those who did not. These were as follows: cholera (Adjusted-OR=6.50, 95%CI,2.30, 18.11),^8^ dysentery ((at households Adjusted-OR=10.52(95%CI,2.81, 39.0), at public gathering (Adjusted-OR=2.92(95%CI,1.24,7.21)),^21^ diarrhoea (Adjusted-OR=2.89, 95%CI,1.33, 6.28),^7^ and Hepatitis E virus (Adjusted-OR=1.90, 95%CI,1.07, 3.38).^6^ See Figure 3 for the forest plot including these odds ratios.

**Figure 3:**
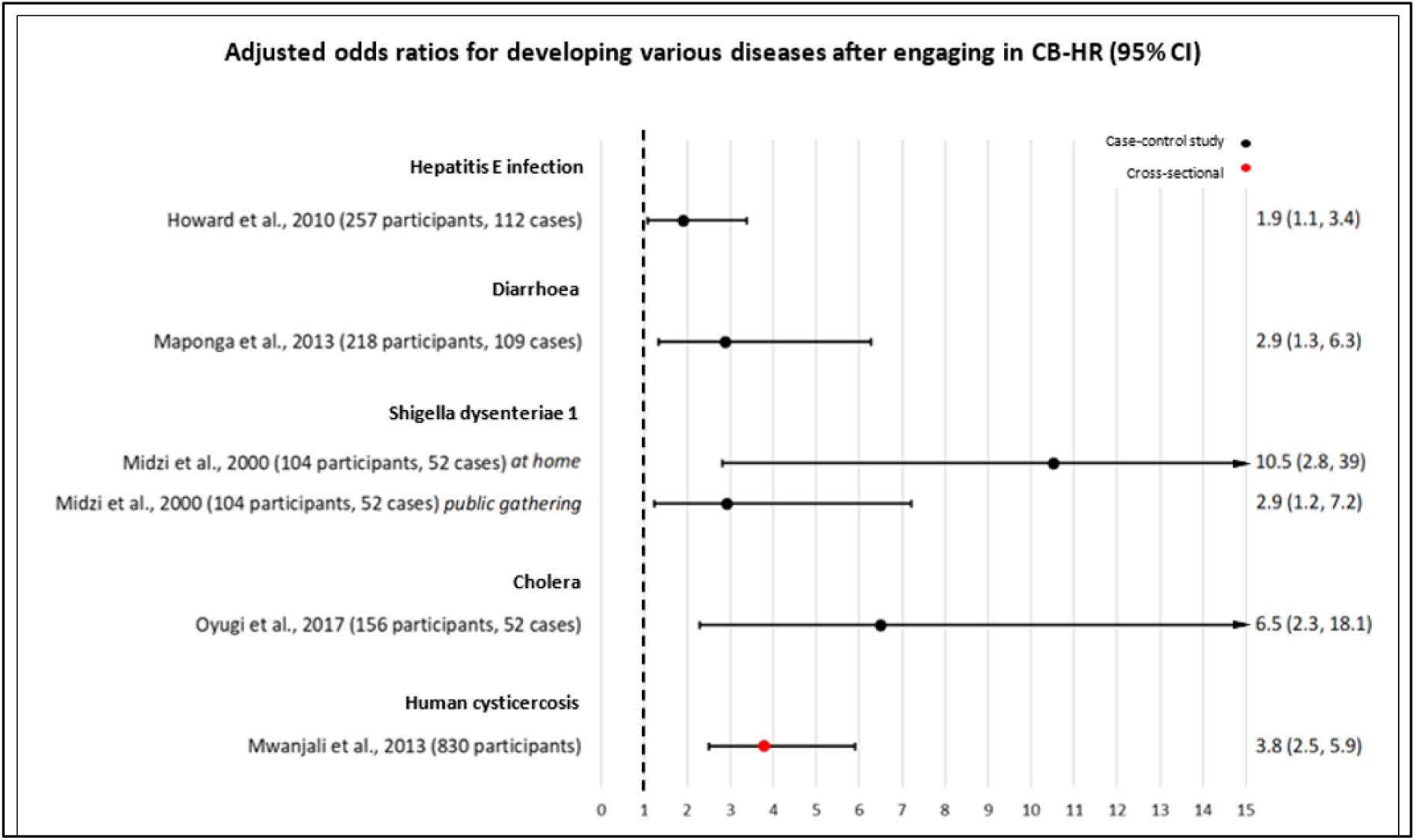
Forest plot of adjusted ORs of disease for those engaging in CB-HR. Adjusted ORs describe odds of disease for those engaging in CB-HR at home versus those who do not practice CB-HR at home. Midzi et al. also estimated an adjusted OR for engaging in CB-HR at public gatherings and this is marked on the plot. Mwanji et al. adjusted for age and whether the participant was a tapeworm carrier. It was not clear which covariates the other studies adjusted for.

Mwanjali et al. ^24^ (a cross-sectional study involving 830 participants ranging from 15-60 years), found an increase in odds of cysticercosis for those who practised CB-HW compared to those who did not for Ag-ELISA seropositivity for *TaeniaSolium* (pork tapeworm) infection and CB-HW (Odds Ratio (OR) = 3.8 (2.5-5.9), p<0.05) (see Figure 3).

### Growth faltering

Rukambile et al. ^22^ in a cohort study found a lower height-for-age z-score in babies who had household exposure to CB-HR practices.

### Microbial content of bowl water and hand-swabs before and after CB-HR

In a school setting, Tetteh-Quarcoo et al. ^10^ using a cross-sectional study design, recorded 54 samples of water and hand-swabs from six preschools. Regarding hand swabs, they investigated several potentially pathogenic bacteria, including Staphylococcus species, Escherichia coli, Citrobacter species, Salmonella species, Klebsiella pneumoniae, and Klebsiella oxytoca. Except for Staphylococcus species which were present both before and after CB-HR; Escherichia coli, Citrobacter species, Salmonella species, Klebsiella pneumoniae, and Klebsiella oxytoca were isolated only after CB-HR. This indicates that the sharing of bowl water during CB-HR led to pathogen transmission and increased the range of bacterial types on hands.

Moabi ^9^ collected 126 samples from one or more bowls at 42 funerals in a cross-sectional study. The samples taken from the bowls before the ritual CB-HR were compared against the safe microbial South African National Standard (SANS – lower levels indicating worst contamination).^25^ Prior to CB-HR, 95.2% of funerals’ water bowls had total aerobic bacteria levels at a safe level when compared to SANS, for coliforms and Escherichia coli. After handwashing, they found that there was significantly greater contamination, with SANS compliance lowered to 45.2% for total aerobic bacteria and for Escherichia coli to 47.6%, indicating a significant possibility that a dangerous level of cross-contamination between participants had taken place.

### Motivations

Among the included studies, seven mentioned reasons for CB-HR practice. However, these were largely author’s speculations without presenting data.^9,15,18,19,21,23^ There were no qualitative in-depth data or explicit objectives to investigate motivations or reasons for the practice. Three main drivers and motivations were speculated: the unpredictability/scarcity of water and washing facilities, cultural/traditional, and/or habitual practices.

Dancesco et al. ^15^ commented that in Côte d’Ivoire, despite the availability of soap, the scarcity of water meant adequate handwashing could not take place. Badowski et al. ^18^ mentioned that in Tanzania the practice of CB-HR was habitual after previous droughts and scarcity of water, compelling households to practice CB-HR to save water. Since then, the practice persisted even through times when water was plentiful. Regarding cultural/traditional beliefs as motivations, Midzi et al. ^21^ and Moabi ^9^ both observed CB-HR being used as part of cultural practices related to cleansing rituals during funerals and other cultural events in South Africa and Zimbabwe.

### Interventions

No studies reported existence of programmes or interventions seeking to reduce or otherwise alter the practice of CB-HR at the household, institutional or other public gatherings level.

## Discussion

This review aims to synthesise published results on the prevalence, risks, and motivations for CB-HR, and existing targeted interventions addressing it. Fifteen studies met the inclusion criteria, eight of which were rated 3 and below on the MMAT 5-point rating criteria, and three mentioned in the text the practice of CB-HR in their population without supportive data. No intervention (randomised trials or other designs) was found. Also, no relevant studies exploring CB-HR outside of SSA were identified, suggesting that CB-HR is predominant in Africa.

Data on prevalence was not available since most studies only examined CB-HR as a risk factor for disease transmission, and did not sample populations randomly (representatively) to assess prevalence of CB-HR. However, studies reported CB-HR occurring in homes, schools and pre-school settings and cultural/social gatherings such as funeral events. This highlights CB-HR as a key area for intervention if hand-transmissible pathogens and diseases are to be prevented and to increase the success of handwashing campaigns.

The most significant findings impelling action towards targeted interventions are the four studies that demonstrated the risk of disease after practising CB-HR.^6–8,21^ They investigated diseases of significant burden, some leading to chronic morbidity and/or mortality (dysentery, cholera, unspecified diarrhoea, hepatitis and cystiserosis).^6,7,8,21^ Tetteh-Quarcoo and colleagues^10^ examined hand-swabs or water from CB-HR and confirmed transmission of pathogens between individuals as a result of CB-HR practice. Similarly, the CB-HR water that had no or little contamination with particular pathogens *before* CB-HR was found to be contaminated with significant concentrations of microbial pathogens *after* CB-HR.^9,10^ Since most CB-HR practice occurs pre-meals, this practice is thus likely to be a contributor to WHO’s estimate of 91 million people in Africa suffering from food-borne diseases, with more than 70% of this burden of foodborne diseases caused by diarrhoeal diseases.^26^

The scarcity of water, habit, and cultural/traditional practices were speculated to be three main reasons for the practice of CB-HR. However, no studies investigated reasons, drivers or motivations for CB-HR. Our recent evidence from WASH studies ongoing in Mali and the Gambia and expert reviews across Africa suggests that CB-HR is still very prevalent in many SSA counties and numerous settings. Our qualitative data from Mali indicate that CB-HW is widely practiced and done mainly to foster social cohesion, that is, improving and maintaining family unity.^27^ There is an urgent need for qualitative or anthropological studies that systematically investigate the reasons and motivations for this practice which has persisted through epidemics, pandemics, and significant handwashing campaigns. Evidence suggests that the years of WASH education have successfully raised the awareness of the population, even in the poorest villages in SSA who are mostly aware that washing with soap and clean water are important hygienic practices before eating.^28^ This review confirms that knowledge alone is not enough for behaviour change, particularly when practices such as CB-HR are culturally and ritualistically entrenched.

It was unexpected that given the high prevalence of CB-HR, we found no targeted interventions reported or evaluated in any country. Our aforementioned research with Water Aid and Save the Children WASH experts across Africa confirms the systematic review findings that CB-HR is highly prevalent in all countries where these NGOs operate and no targeted interventions were known to these experts.^29^

Any CB-HR interventions development should be sensitive to cultural beliefs and practices. Formative research tailored by region would ensure interventions are culturally sensitive and address motivators and beliefs underlying CB-HR.^5,30^

## Strengths and limitations

Strengths of our systematic review include searches in numerous databases; manual screening of the reference lists of all included articles; the screening of grey literature, not including any date limit, English and French languages included on searches, and contacting a range of experts in retrieving unpublished reports on CB-HW. Additionally, the use of two independent reviewers during the screening phase, data extraction and quality assessment of this review plus a third independent arbiter enhanced the internal reliability and consistency of included articles.

In terms of limitations, due to the heterogeneity of identified studies with a range of designs and methodological differences, a meta-analysis could not be performed. Thus, conclusive combined statistical results could not be generated; instead, a narrative synthesis was conducted to analyse the results. Further, this review found only observational, and no experimental studies. Due to the lack of experimental studies, our confidence in the conclusions drawn from the included studies is limited, particularly for the findings exploring the disease risks of CB-HR. However, the apparent lack of interventional studies regarding CB-HR offers an opportunity for future research in this critical WASH area.

## Conclusions

This review indicates that CB-HR is widespread in Sub-Saharan Africa, and has a high risk of pathogen and disease transmission, even to vulnerable young children in households or at pre-schools. Evidence links the increased risk for disease transmission through CB-HR to outbreaks of various significant gastro-intestinal diseases such as cholera, dysentery, and hepatitis infection. Although evidence is clear about the widespread of CB-HR across SSA, given the scanty and poorly reported literature, more research is needed to provide a better understanding of the spread and prevalence of this practice across a range of settings including households, schools, workplaces and social gatherings (e.g. funerals).

In spite of evidence demonstrating CB-HR to be an important and widespread risky hygienic practice in Africa, we have found no research into the drivers and motivators for the practice or evidence of the development and evaluations of interventions to reduce the dangers inherent in CB-HR. Therefore, we recommend anthropological, sociological and qualitative studies to understand motivators and drivers for CB-HR practices, and barriers and facilitators for behaviour change at the household, community or institutional level where CB-HR continues.

Building on these studies, interventions to modify CB-HR should be developed and evaluated with families, schools, community/religious leaders to enable behaviour change at the community-level. Engaging policy-makers early in the co-development of policies will be critical to removing barriers to accessing water and WASH facilities and promoting the development of evidence-based culturally acceptable interventions.

## Data Availability

All data produced in the present work are contained in the manuscript

## Abbreviations

CB-HR: Communal Bowl-Hand Rinsing
MMAT: Mixed Methods Appraisal Tool
ECD: Early Childhood Development
HEV: Hepatitis E virus
E.Coli: Escherichia coli
WHO: World Health Organisation

## Authors’ contributions

EAA and SMH conceived the idea and developed the initial plan. AR and AB did the screening. Quality checks and data extraction were done by EAA, SMH and JAA. KS provided statistical expertise and produced figures. RL, OT, MY, CS and YD provided technical support and advice. All authors read and approved the final version.

## Research Ethics statement

Non-Applicable

