## Supplementary File for "Good Intentions but bad outcomes – Communal-bowl hand-rinsing before meals transmits pathogens and diseases: a systematic review"

**Supplementary materials**

**Appendix A: Search strategy**

| 1. | communal or common or shared or together or joint or mutual or pooled or community or general or collective |
| --- | --- |
| 2. | hand adj3 (wash* OR clean* OR sanitis* OR sanitiz* OR disinfect* OR rinse OR hygiene OR swab) |
| 3. | 1 AND 2 |

**Appendix B: Eligibility Criterion**

| **Criteria** | **Inclusion** | **Exclusion** |
| --- | --- | --- |
| Population | Study participants are unspecified, all populations are included. | Studies examining handwashing without focusing on communal handwashing. |
| Practice | Studies examining communal handwashing (or the practice of together washing in a bowl). | Studies that only examine food hygiene (including infant weaning food). |
| Outcome measures | Studies where the outcomes assessed include microbial load of communal water or microbial content of the hands of people practicing communal handwashing, the risk of the disease, the prevalence, and motivations for C-HW | Studies that only examine household hygienic conditions such as the kitchen. |
| Language | Studies published in English or French. | Studies were conducted using animal or plant cell lines (in vitro or in vivo). |

**Appendix C – Summary of Study Quality using MMAT**

|  |  | **MMAT Questions** | | | | | | |  |
| --- | --- | --- | --- | --- | --- | --- | --- | --- | --- |
|  | **Author (quantitative descriptive)** | **S1-Are there clear research questions?** | **S2- Do the collected data allow to address the research questions?** | **X.1- Is the sampling strategy relevant to address the research question?** | **X.2- Is the sample representative of the target population?** | **X.3-Are the measurements appropriate?** | **X.4-Is the risk of nonresponse bias low?** | **X.5-Is the statistical analysis appropriate to answer the research question?** | **Rating (0* lowest and 5*highest)** |
| 1 | Appiah-Brempong et al 2018 | **Yes** | **yes** | **yes** | **yes** | **yes** | **yes** | **Yes** | **5*** |
| 2 | Dancesco et al 2005 | No | yes | yes | yes | no | no | no | 2* |
| 3 | Ehiri et al., 2001 | yes | Yes | yes | yes | yes | not sure | yes | 4* |
| 4 | Howard et al., 2010 | yes | yes | yes | no | yes | not sure | yes | 3* |
| 5 | Maponga et al., 2011 | yes | yes | yes | yes | yes | not sure | yes | 4* |
| 6 | Melariri et al., 2019 | yes | yes | Yes | no | yes | not sure | yes | 3* |
| 7 | midzi et al 2000 | yes | yes | yes | no | yes | not sure | yes | 4* |
| 8 | Moabi, 2016 | yes | yes | yes | No | yes | No | yes | 2* |
| 9 | Mwanjali et al., 2013 | yes | yes | yes | yes | yes | yes | yes | 5* |
| 10 | Mwapasa et al., 2022 | yes | yes | yes | ? | yes | not sure | yes | 3* |
| 11 | Oyugi et al., 2017 | yes | yes | yes | yes | yes | not sure | yes | 4* |
| 12 | Rukambile et al., 2020 | yes | yes | yes | yes | yes | not sure | yes | 4* |
| 13 | Schmitt et al 1997 | yes | yes | yes | no | no | no | no | 1* |
| 14 | Tetteh-Quarcoo et al., 2016 | yes | yes | no | no | yes | not sure | yes | 2* |
| Qualitative study | | | | | | | | | |
|  | **Author (qualitative)** | **S1-Are there clear research questions?** | **S2- Do the collected data allow to address the research questions?** | 1.1. Is the qualitative approach appropriate to answer the research question? | 1.2. Are the qualitative data collection methods adequate to address the research question? | 1.3. Are the findings adequately derived from the data? | 1.4. Is the interpretation of results sufficiently substantiated by data? | 1.5. Is there coherence between qualitative data sources, collection, analysis and interpretation? | **Total** |
| 15 | Badowski et 2011 | yes | not sure | yes | no | yes | not sure | yes | 3* |
